# Soluble immune checkpoints are dysregulated in patients with sickle cell disease and correlate with inflammatory mediators, autoantibodies, immune cell profiles, and clinical outcomes

**DOI:** 10.1101/2025.02.27.25322979

**Authors:** Wei Li, Andrew Q Pucka, Lina Houran, Xiaoqing Huang, Candice Debats, Brandon A Reyes, Andrew RW O’Brien, Qigui Yu, Ying Wang

## Abstract

**Background:** Sickle cell disease (SCD) is a chronic condition characterized by inflammation, immune dysregulation, and debilitating pain.

**Aim:** This study investigates soluble immune checkpoints (sICPs) and their associations with inflammatory mediators, immune cell profiles, autoantibodies, and clinical outcomes in SCD.

**Method:** Peripheral blood samples from 50 SCD patients and 40 demographic-matched healthy controls (HCs) were analyzed for 37 sICPs, 80 inflammatory mediators, and 18 autoantibodies using multiplex assays, alongside immune cell profiles via flow cytometry. Pain and quality of life (QoL) were assessed through patient-reported outcome measures (PROMs).

**Results:** Twenty-three sICPs, including arginase-1, BTLA, CD27, CD28, CD47, CD80, CD96, CD134, CD137, CD152, GITR, HVEM, IDO, LAG-3, MICA, MICB, Nectin-2, PD-1, Siglec-7, Siglec-9, TIM-3, TIMD-4, and VISTA, were significantly elevated in SCD patients compared to HCs. These sICPs correlated with multiple proinflammatory mediators (e.g., IL-18), autoantibodies (e.g., MPO), and immune cell activation markers (e.g., CD38/HLA-DR on CD8 T cells). Notably, CD28, CD152, HVEM, and VISTA were strongly associated with systemic inflammation and immune cell activation, while BTLA, LAG-3, PD-1, and CD80 correlated with pain and anxiety scores and QoL.

**Conclusion:** This study highlights complex interactions between sICPs, immune activation, inflammation, and clinical outcomes in SCD, underscoring their potential as biomarkers or therapeutic targets to alleviate inflammation and improve QoL in this challenging clinical population.

## 1. Introduction

Sickle cell disease (SCD) is a complex, lifelong condition characterized by systemic involvement and significant complications, including acute and chronic pain, anemia, stroke, pulmonary hypertension, and progressive organ damage (1, 2). Clinically, SCD alternates between two distinct states: the steady-state (StSt) phase, often presenting with mild symptoms related to chronic hemolysis and ongoing discomfort, and the acute vaso-occlusive crisis (VOC) phase, marked by severe pain episodes and potential life-threatening complications such as acute chest syndrome and stroke due to blood vessel blockages by sickle-shaped red blood cells (RBCs) (3–5). Central to the pathology of SCD is a state of chronic inflammation and immune dysregulation, which underpins the disease’s clinical manifestations and contributes to its progression.

Chronic inflammation in SCD patients is driven by various mechanisms, including hemolysis, ischemia-reperfusion injury, oxidative stress, and immune dysregulation. Hemolysis releases hemoglobin and heme into circulation, triggering the activation of immune cells through NF-κB and TLR4 pathways, resulting in elevated levels of inflammatory cytokines such as IL-1β, IL-6, and TNF-α (6–8). Additionally, the ischemic and hypoxic conditions caused by vascular occlusion activate endothelial cells, promoting the expression of adhesion molecules and the recruitment of leukocytes, which further amplify the inflammatory cascade (9, 10). Autoimmune processes also play a role, with the production of autoantibodies driven by aberrant B-cell activation contributing to immune complex formation and tissue damage (11, 12). These sustained inflammatory responses are compounded by the oxidative stress inherent to SCD, further exacerbating endothelial dysfunction and immune cell activation.

Despite advances in understanding SCD pathophysiology, gaps remain in identifying the full spectrum of immune regulatory mechanisms involved in the disease. Recent advances have identified soluble immune checkpoints (sICPs) as critical modulators of immune regulation and inflammation (13, 14). Soluble forms of immune checkpoint molecules, generated via protease-mediated shedding or alternative mRNA splicing, act as circulating regulators of immune responses. They play diverse roles in maintaining immune homeostasis, interacting with inflammatory pathways, and acting as biomarkers of disease severity or therapeutic response. While sICPs have been recently studied in conditions such as cancer, chronic viral infections, and autoimmune diseases, their role in SCD remains unexplored.

Given that chronic inflammation and immune dysregulation are prominent in SCD patients, studying sICPs could provide valuable insights into the disease’s inflammation and immune landscape. These molecules may represent novel biomarkers of disease severity or therapeutic targets, offering a new avenue for understanding and managing SCD. This study seeks to address this gap by utilizing baseline clinical samples from our clinical trial (NCT05045820) to investigate the levels and roles of sICPs in SCD for the first time. Alongside profiling inflammatory mediators, autoantibodies, and immune cell profiles, we aim to elucidate the associations between sICPs, inflammation, immune dysregulation, and clinical outcomes, including VOCs, pain severity, and quality of life (QoL) measures. By integrating this analysis, we provide insights into mechanisms underlying SCD pathophysiology and identify potential targets for innovative therapeutic interventions.

## 2. Materials and Methods

### 2.1 Study Participants

This study is part of our ongoing randomized clinical trial in SCD (ClinicalTrials.gov, NCT05045820) (15). Eligible participants aged 14 to 80 were individuals diagnosed with SCD who experienced chronic pain within the last 6 months and/or at least one vaso-occlusive crisis (VOC) in the past 12 months. Participants also met the following inclusion criteria: no recent changes in stimulant medication, willingness to continue ongoing treatments, and agreement to avoid new medications or pain management methods during the study. Major exclusion criteria included confirmed or suspected COVID-19, recent or ongoing acupuncture for pain management (within the last 6 months), presence of autoimmune or inflammatory diseases, and blood transfusion within 90 days before enrollment. Age-, gender-, and ethnicity-matched healthy controls (HCs) without SCD were also recruited. Comprehensive criteria are available at ClinicalTrials.gov (NCT05045820).

Peripheral blood was collected from 50 SCD participants at steady-state (StSt) (58% female; median age: 31 years) and 40 HCs (60% female; median age: 33.5 years). All participants were Black/African American. Blood samples were collected between July 2021 and June 2024 from Indiana University Health hospitals, the Indiana Hemophilia & Thrombosis Center, community hospitals, and other external medical facilities. Plasma was separated and stored at -80°C, while peripheral blood mononuclear cells (PBMCs) were cryopreserved in liquid nitrogen or used directly. Detailed demographic, clinical, and laboratory characteristics are summarized in Table 1.

**Table 1.** Demographics and clinical characteristics of SCD patients.

| Parameters | HCs (n=40) | SCD (n=50) | p value |
| --- | --- | --- | --- |
| **Demographics** |  |  |  |
| Age (years) | 33.5 (20.5-48.0) | 31 (23.5-42.3) | 0.76 |
| Gender (% females) | 24 (60%) | 29 (58%) | 0.86 |
| SS/Sb0/SC/Sb+ thalassemia/$\alpha$ thalassemia (n/n/n/n) | -- | 29/3/14/3/1 | |
| Body mass index (kg/m<sup>2</sup>) | 27.0 (23.8-33.9) | 23.8 (20.8-28.4) | 0.001 |
| Infarction | -- | 5 (10%) |  |
| **Disease-Modifying Therapies** |  |  |  |
| Hydroxyurea | -- | 24 (48%) |  |
| Chronic Transfusion Therapy | -- | 6 (12%) |  |

**Note:** Data are shown as median and interquartile range or number and percentage. **SCD**, sickle cell disease; **HC**, healthy controls. The Mann-Whitney test was used to compare differences between SCD patients and HCs for continuous variables. $\chi^2$ test was used for comparison of gender distribution between the two groups. $p < 0.05$ was considered significant.

### 2.2 Patient-Reported Outcome Measures (PROMs)

PROMs were collected using validated tools as used in our previous published work (15). The PainDETECT Questionnaire assessed neuropathic pain (higher scores indicated higher pain) (16, 17), while PROMIS-29 assessed pain intensity, interference, and physical dysfunction (18). The recency and frequency of VOCs were assessed by the Adult Sickle Cell Quality-of-Life Measurement Information System (ASCQ-Me) (19). Depression and anxiety were evaluated with the Hospital Anxiety and Depression Scale (HADS), and pain-related quality of life (QoL) was measured using the Pediatric Quality of Life Inventory (PedsQL, both peds and adult versions) (20). The Fibromyalgia Survey Questionnaire (FSQ), which consists of the Widespread Pain Index, was utilized as a surrogate measure of nociplastic pain (21). Results from these assessments, including significant differences between SCD and HC groups, are presented in Table 2.

**Table 2.** Patient-reported outcome measures (PROMs)

| PROMs | HC (n=31-40) | SCD (n=37-48) | p value |
| --- | --- | --- | --- |
| PainDetect_Total score | 7 (7-7) | 19 (13-24) | <0.0001 |
| BPI_Pain Interference Score | 0 (0-0) | 3.7 (0.7-3.7) | <0.0001 |
| FSQ_Widespread Pain Index | 0 (0-0) | 6 (2-8) | <0.0001 |
| PROMIS-29_Physical Function Score | 4 (4-4) | 8 (5-12) | <0.0001 |
| PROMIS-29_Pain Intensity Score | 0 (0-1) | 5 (4-6) | <0.0001 |
| HADS_Depression Score | 1 (0-3) | 6 (2-8) | 0.0001 |
| HADS_Anxiety Score | 2 (1-4) | 6 (3-8) | <0.0001 |
| ASCQ-Me_Pain Episode Frequency Score | N/A | 8 (6-9) |  |
| Number of VOCs in preceding 12 months | N/A | 5 (2-7) |  |
| PedsQL Total Score | N/A | 56 (44-56) | <0.0001 |

**Note:** Data are shown as median and interquartile range. **HC**, healthy control; **SCD**, sickle cell disease; **BPI**, Brief Pain Inventory; **FSQ**, Fibromyalgia Survey Questionnaire; **PROMIS**, Patient-Reported Outcomes Measurement Information System; **HADS**, Hospital Anxiety and Depression Scale; **ASCQ-Me**, Adult Sickle Cell Quality of Life Measurement Information System; **VOC**, vaso-occlusive crisis; **PedsQL**, Pediatric Quality of Life Inventory. The Mann-Whitney test was used to compare differences between SCD patients and HCs. $p < 0.05$ was considered significant.

### 2.3 Multiplex immunoassays

The ProcartaPlex Human Immune Checkpoint Panel 37plex (Cat. #: EPX370-15846-901, Invitrogen, Carlsbad, CA) was used to quantify plasma concentrations of thirty-seven soluble immune checkpoints (sICPs), including twenty inhibitory ICPs (Arginase-1, B7-H6, BTLA, CD47, CD48, CD73, CD152, CD276, HVEM, IDO, LAG-3, PD-1, PD-L1, PD-L2, PVR, S100A8/A9, Siglec-7, Siglec-9, TIM-3, and VISTA) and seventeen stimulatory ICPs (CD27, CD28, CD80, CD96, CD134, CD137, E-Cadherin, GITR, ICOSL, MICA, MICB, Nectin-2, Perforin, TIMD-4, ULBP-1, ULBP-3, and ULBP-4). ProcartaPlex Human Immune Response Panel 80plex (Cat. #: EPX800-10080-901, Invitrogen, Carlsbad, CA) was used to quantify plasma concentrations of eighty inflammatory mediators, including twenty-seven inflammatory cytokines (IFN-α, IFN-γ, IL- 1α, IL-1β, IL-2, IL-3, IL-4, IL-5, IL-6, IL-9, IL-10, IL-12p70, IL-13, IL-15, IL-16, IL-17A, IL-18, IL-21, IL-22, IL-23, IL-27, IL-31, IL-37, MIF, TNF-β, TNF-α, and TSLP), twenty-seven chemokines (BLC, CCL1, CCL17, CCL21, CCL23, CCL25, CXCL6, ENA-78, Eotaxin, Eotaxin-2, Eotaxin-3, Fractalkine, Gro-α, IL-8, IP-10, I-TAC, MCP-1, MCP-2, MCP-3, MCP-4, MDC, MIG, MIP-1α, MIP-1β, MIP-2α, MIP-3α, and MIP-3β), twelve growth factors (bNGF, FGF-2, G-CSF, GM-CSF, HGF, IL-7, IL-20, IL-34, LIF, M-CSF, SCF, and VEGF-A), twelve soluble receptors/proteins (APRIL, BAFF, CD30, CD40L, Gal-3, IL-2R, MMP-1, PTX3, TNF-RII, TRAIL, TREM-1, and TWEAK), and two serine proteases (Granzyme A and Granzyme B). The MILLIPLEX MAP Human Autoimmune Autoantibody Panel (Cat. #: HAIAB-10K, MilliporeSigma, Burlington, MA) was used to quantify plasma concentrations of eighteen anti-human autoantibodies, including anti-C1q, anti-Centromere Protein A (CENP-A), anti-Centromere Protein B (CENP-B), anti-β 2-glycoprotein, anti-Ku, anti-Mi-2, anti-myeloperoxidase, anti-proliferating cell nuclear antigen A (PCNA), anti-PM/Scl-100, anti-proteinase 3, anti-Ribosomal P, anti-Ribonucleoprotein (RNP), anti-RNP/Smith (RNP/Sm), anti-Scl-70, anti-Sm, anti-Sjögren′s Syndrome-related antigen B/La (SSB/La), anti-anti-Sjögren′s Syndrome-related antigen A/Ro52 kDa (SSA/Ro52), and anti-Sjögren′s Syndrome-related antigen A/Ro60 kDa (SSA/Ro60). The beads were read on a BioPlex 200 system (Bio-Rad, Hercules, CA). The standards at 4-fold serial dilutions were run on each plate in duplicate and used to calculate the concentrations of sICPs and inflammatory mediators using the Bio-Plex Manager Software (Bio-Rad, Hercules, CA) as previously reported (15, 22). Plasma samples were diluted 100-fold for the autoantibody multiplex assay, and the levels of the autoantibodies were reported as mean fluorescence intensity (MFI) after background MFI subtraction.

### 2.4 Flow cytometry

Frozen PBMCs were incubated with fixable viability dye (Thermo Fisher Scientific, Cincinnati, OH, USA) to exclude dead cells for analysis. Cells were then stained with fluorochrome-conjugated antibodies against cell lineage markers (CD3, CD4, CD14, CD16, CD19, CD56, CD161, and MR1-tetramer loaded with the ligand 5-OP-RU) and three immune cell activation markers (CD38, CD69, and HLA-DR) and two membrane ICP (mICPs)/exhaustion markers (PD- 1 and TIM-3). Isotype control antibodies were used to set gates for positive expression for the activation markers and mICPs. All antibodies were purchased from BioLegend (San Diego, CA, USA). The MR1 tetramers were produced by the NIH Tetramer Core Facility as permitted to be distributed by the University of Melbourne. Stained cells were acquired using a BD LSRFortessa flow cytometer (BD Biosciences, San Jose, CA). Flow data were analyzed using FlowJo v10 software (Tree Star, San Carlos, CA).

### 2.5 Statistical analysis

Statistical analysis was performed using GraphPad Prism 10 and SPSS 29. Data were expressed as median and interquartile range. Differences between 2 groups were calculated using the Mann-Whitney test for continuous variables. χ 2 test was used for comparison between 2 groups for gender distribution. Adjusted *p* values were calculated for the 37-plex, 80- plex, 18-plex analytes, and immune cell phenotypes using the Holm-Šídák correction for multiple comparisons). Inflammatory mediators that were heightened in the SCD participants were used for subsequent Spearman correlation analyses using age, gender, SCD genotype, hydroxyurea (HU) treatment, BMI, and chronic transfusions as covariables. *p* < 0.05 was considered statistically significant.

## 3. Results

### 3.1 Characteristics of the SCD study cohort: demographics and patient-reported outcomes

The demographic and clinical characteristics of 50 SCD participants, alongside 40 healthy controls (HCs), are summarized in Table 1. There were no significant differences between SCD participants and HCs in terms of age (median: 31 vs. 33.5 years; *p* = 0.76) or gender distribution (58% vs. 60% females; *p* = 0.85). Body mass index (BMI) was significantly lower in the SCD group compared to HCs (median: 23.8 vs. 27.0 kg/m²; *p* = 0.001). Five SCD participants (10%) had a history of infarction. Among SCD participants, 48% were on HU therapy, and 12% received chronic transfusion therapy.

Patient-reported outcome measures (PROMs) indicated significantly greater pain burden and impaired quality of life (QoL) in SCD participants compared to HCs, as detailed in Table 2. SCD participants reported higher PainDETECT scores (median: 19 vs. 7; *p* < 0.0001), greater pain interference (median Brief Pain Inventory score: 3.7 vs. 0; *p* < 0.0001), and more widespread pain (FSQ Widespread Pain Index: 6 vs. 0; *p* < 0.0001). Physical functioning was significantly worse among SCD participants (PROMIS-29 Physical Function Score: median 8 vs. 4; *p* < 0.0001), while depression and anxiety levels were elevated (HADS Depression: 6 vs. 1; *p* = 0.0001; HADS Anxiety: 6 vs. 2; *p* < 0.0001). SCD participants reported multiple VOCs in the past 12 months (median: 5 events) and lower pain-related QoL (PedsQL Total Score: 56 vs. 100; *p* < 0.0001).

### 3.2 Circulating sICPs were highly dysregulated in SCD patients

All sICPs except four (CD48, ULBP-1, ULBP-3, and ULBP-4) were detected in the plasma samples from both the SCD patients and HCs. SCD patients exhibited significantly dysregulated levels of multiple sICPs compared to HCs (Table 3). Among the twenty inhibitory sICPs examined, twelve of them, including arginase-1, BTLA, CD47, CTLA-4 (CD152), HVEM, IDO, LAG-3, PD-1, Siglec-7, Siglec-9, TIM-3, and VISTA, were elevated in SCD patients. After adjusting for multiple comparisons, seven (CD47, CTLA-4 (CD152), HVEM, LAG-3, Siglec-7, TIM-3, and VISTA) remained significantly upregulated. Among the seventeen stimulatory sICPs examined, eleven of them, including CD27, CD28, CD80, CD96, CD134 (OX40), CD137 (4- 1BB), GITR, MICA, MICB, Nectin-2, and TIMD-4, were markedly elevated in SCD patients, and all except CD96 and GITR remained significantly different after multiple comparison adjustment (Table 3). These findings highlight a dysregulated circulating immune profile.

**Table 3.**
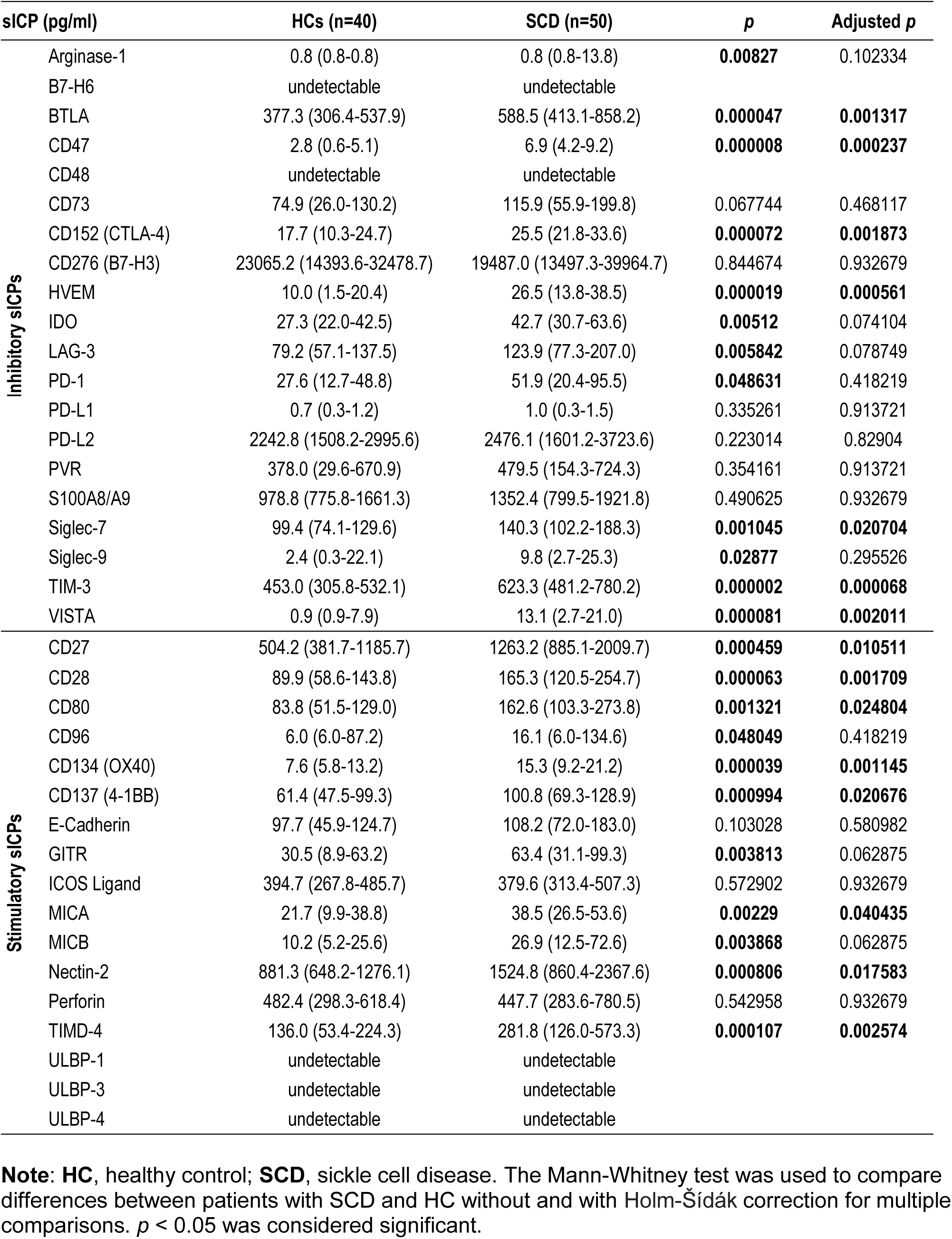
Plasma levels of sICPs in SCD patients versus HCs.

The adjusted analysis identified a subset of sICPs that were robustly increased, including BTLA, CD47, CD152, HVEM, TIM-3, VISTA, CD28, CD134, and TIMD-4, indicating their potential as biomarkers for immune dysfunction in SCD (Table 3) Interestingly, TIM-3, an inhibitory sICP linked to immune exhaustion, was among the most significantly elevated markers in SCD participants, emphasizing its role in immune dysregulation. The stimulatory checkpoint CD28, critical for T cell activation, showed pronounced upregulation in SCD participants. These data underscore the complex interplay between inhibitory and stimulatory sICPs in the immunopathogenesis of SCD, with a distinct shift towards heightened immune activation and exhaustion.

### 3.3 Comprehensive analysis revealed correlations of sICPs with inflammatory mediators and autoantibodies in SCD patients

To explore the interplay between immune regulation and inflammation in SCD patients, we performed a Spearman correlation analysis of elevated sICPs with dysregulated inflammatory mediators (Table S1) and autoantibodies (Table S2) (15). Notably, the associations varied depending on whether covariables (age, gender, SCD genotype, HU use, BMI, chronic transfusion) were included in the analysis. For simplicity, we only reported correlation results from multivariant analysis. Our analysis revealed significant associations of plasma levels of sICPs with inflammatory mediators as depicted in a heatmap (Fig. 1), where stronger correlations are represented by higher intensity of color gradients (coefficient) and number of stars (*p* value). Specifically, twenty sICPs, including BTLA, CD47, CD152, HVEM, IDO, LAG-3, PD-1, Siglec-7, TIM-3, VISTA, CD27, CD28, CD80, CD134, CD137, GITR, MICA, MICB, Nectin-2, and TIMD-4, demonstrated positive correlations with various proinflammatory mediators, including multiple inflammatory cytokines (such as IFN-γ, IL-4, IL-6, IL-18, and TNF-α), chemokines (such as CCL21, IL-8, and MIP-1β), growth factors (such as IL-20, IL-34, and VEGF-A). A subset of the sICPs also positively correlated with two soluble receptors (PTX3 and TREM-1) and two serine proteases (granzyme A and granzyme B). Several upregulated sICPs, such as CD152, HVEM, LAG-3, VISTA, CD28, CD80, GITR, and MICA, were highly associated with many inflammatory mediators. These results indicate a complex network of interactions between sICPs and inflammatory mediators, underscoring the potential role of specific sICPs in driving or modulating inflammation in SCD patients.

**Figure 1.**
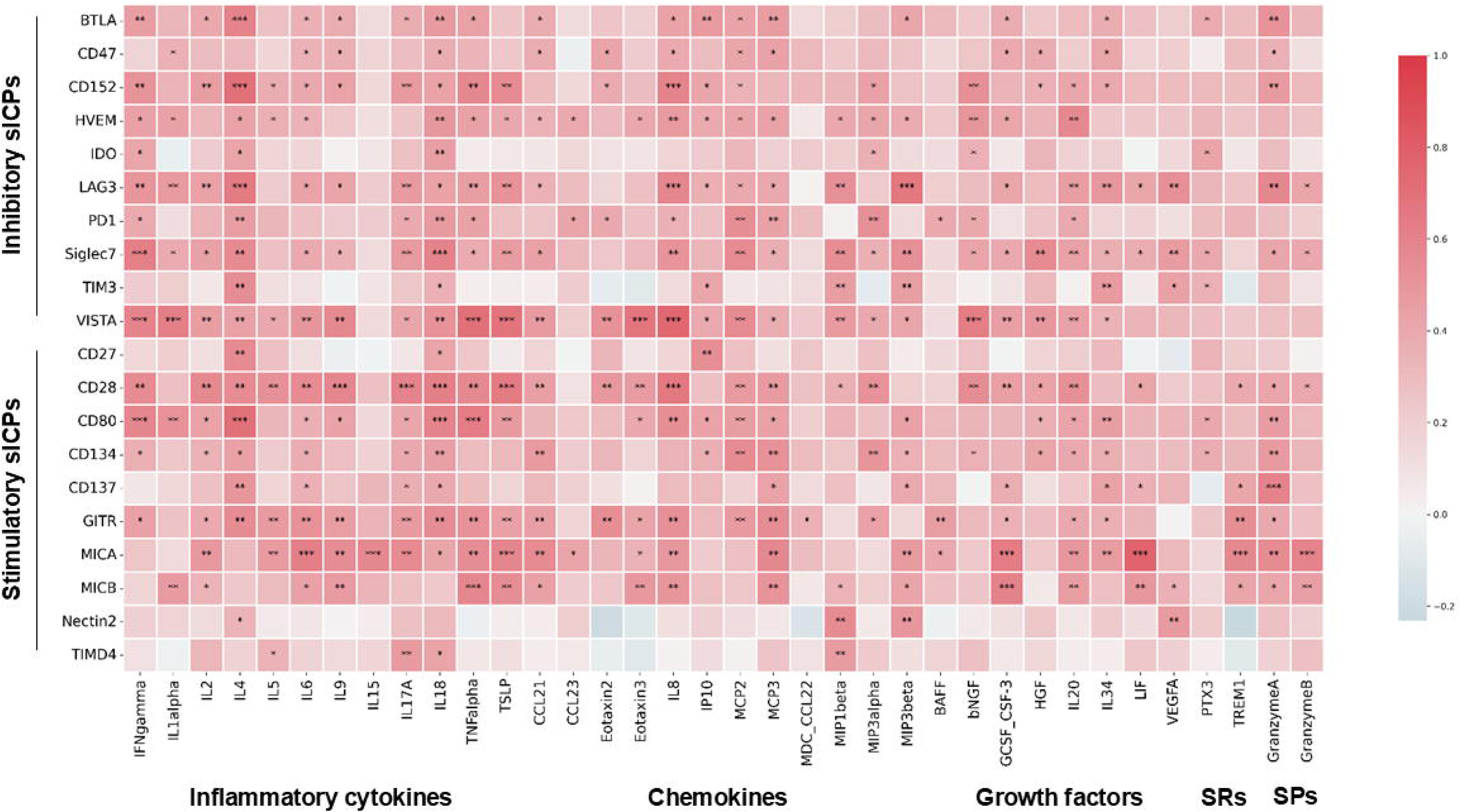
Correlations between altered plasma levels of sICPs and inflammatory mediators in participants with sickle cell disease. Heatmap showing Spearman correlation coefficient (color) and p values (stars). Covariables include age, gender, hydroxyurea, body mass index, and chronic transfusion. sICPs, soluble immune checkpoints; SCD, sickle cell disease; SPs, serine proteases; SRs, soluble receptors. \**p* < 0.05; \*\**p* < 0.01; \*\*\**p* < 0.001.

Similarly, we also performed a Spearman correlation analysis of the dysregulated sICPs with eight autoantibodies that were significantly upregulated in SCD patients relative to HCs (Ku, PM/Scl-100, ribosomal P, RNP/Sm, SSB/La, SSA/Ro60, myeloperoxidase, and proteinase 3) and one that trended higher (Smith antigens, Sm) (Table S2). This analysis revealed significant associations between plasma levels of three sICPs and specific autoantibodies, represented by coefficient values and significance levels. The inhibitory sICPs CD47 and Siglec-7 demonstrated significant positive correlations with autoantibodies against myeloperoxidase and ribosomal P protein and Sm, respectively, while the stimulatory sICP CD80 exhibited significant positive correlations with the Ku and Sm autoantibodies (Table 4). These findings reveal intricate networks of interactions between sICPs and autoantibodies, emphasizing the dual roles of these molecules in both promoting and potentially mitigating autoimmunity in SCD patients.

**Table 4.** Correlations between altered plasma levels of sICPs and autoantibodies.

| sICPs | Ku | Rib-P | Sm | MPO |
| --- | --- | --- | --- | --- |
| **Inhibitory sICPs** |  |  |  |  |
| CD47 |  |  |  | 0.41* |
| Siglec-7 |  | 0.45* | 0.44* |  |
| **Stimulatory sICPs** |  |  |  |  |
| CD80 | 0.40* |  | 0.46* |  |

**Note:** **sICPs**, soluble immune checkpoints. Anti-Ku, anti-Ribosomal P (**Rib-P**), and anti-Smith (**Sm**) autoantibodies are categorized as anti-nuclear autoantibodies (**ANAs**), while anti-myeloperoxidase (**MPO**) falls under anti-non-nuclear autoantibodies. The Spearman correlation test was performed, generating coefficient values (*r*) and statistical significance levels (*p*). Symbols indicating significance: \**p* < 0.05. *p* < 0.05 was considered statistically significant.

### 3.4 Comprehensive analysis revealed significant correlations of sICPs with alterations of immune cell profiles in SCD patients

We first compared immune cell profiles in SCD patients and HCs by multiparametric flow cytometry (Fig. S1). PBMCs were immunophenotyped with cell lineage markers (CD3, CD4, CD14, CD16, CD19, CD56, CD161, and MR1-tetramer loaded with the ligand 5-OP-RU), three immune cell activation markers (CD38, CD69, and HLA-DR) and two membrane ICP/exhaustion markers (PD-1 and TIM-3). As shown in Table S3, the immunophenotypes of both innate (monocytes and NK cells) adaptive immune cells (various subsets of T cells, including MAIT, NKT, CD4 and CD8 T cells) were altered in SCD patients. In particular, NK cells and all subsets of T cells expressed higher levels of the cell activation marker CD69. NKT cells, CD8 T cells, and B cells also had higher levels of two other cell activation markers, CD38 and HLA-DR. On the other hand, the classic monocytes downregulated the inhibitory mICP TIM-3 and the monocyte lineage marker CD14, and the B cells trended to have lower levels of TIM-3 expression (*p* = 0.07). Additionally, the frequency of NK cells trended higher (*p* = 0.06), while that of T cells was significantly lower in SCD patients. These results indicated dysregulation and hyperactivation of the immune system in SCD patients.

To investigate the interplay between dysregulated sICPs and altered immune cell profiles in SCD patients, Spearman correlation analyses were performed with covariables, which included age, gender, SCD genotype, HU use, BMI, and chronic transfusion status (Table 5). Both inhibitory and stimulatory sICPs demonstrated significant associations with phenotypes of various immune cells, including subsets of monocytes, NK cells, T cells, MAIT cells, NKT cells, activated T cells, and B cells. Specifically, nine sICPs (CD47, CD152, IDO, LAG-3, TIM-3, CD28, CD80, GITR, and Nectin-2) displayed positive correlations with phenotypes of multiple immune cell types. Conversely, four sICPs (HVEM, Siglec-7, VISTA, and TIMD-4) showed both positive and negative correlations with immune cell phenotypes. Notably, five sICPs (HVEM, Siglec-7, VISTA, MICB, and TIMD-4) exhibited significant negative correlations with the decreased percentage of T cells, while no sICPs were positively correlated with T cell percentages. Furthermore, CD152, HVEM, VISTA, and CD28 correlated with multiple aspects of immune cell phenotypes. These findings highlight the intricate and multifaceted interactions between sICPs and immune cell profiles in SCD patients, underscoring the need for further investigation to elucidate their immunological and clinical implications.

**Table 5.**
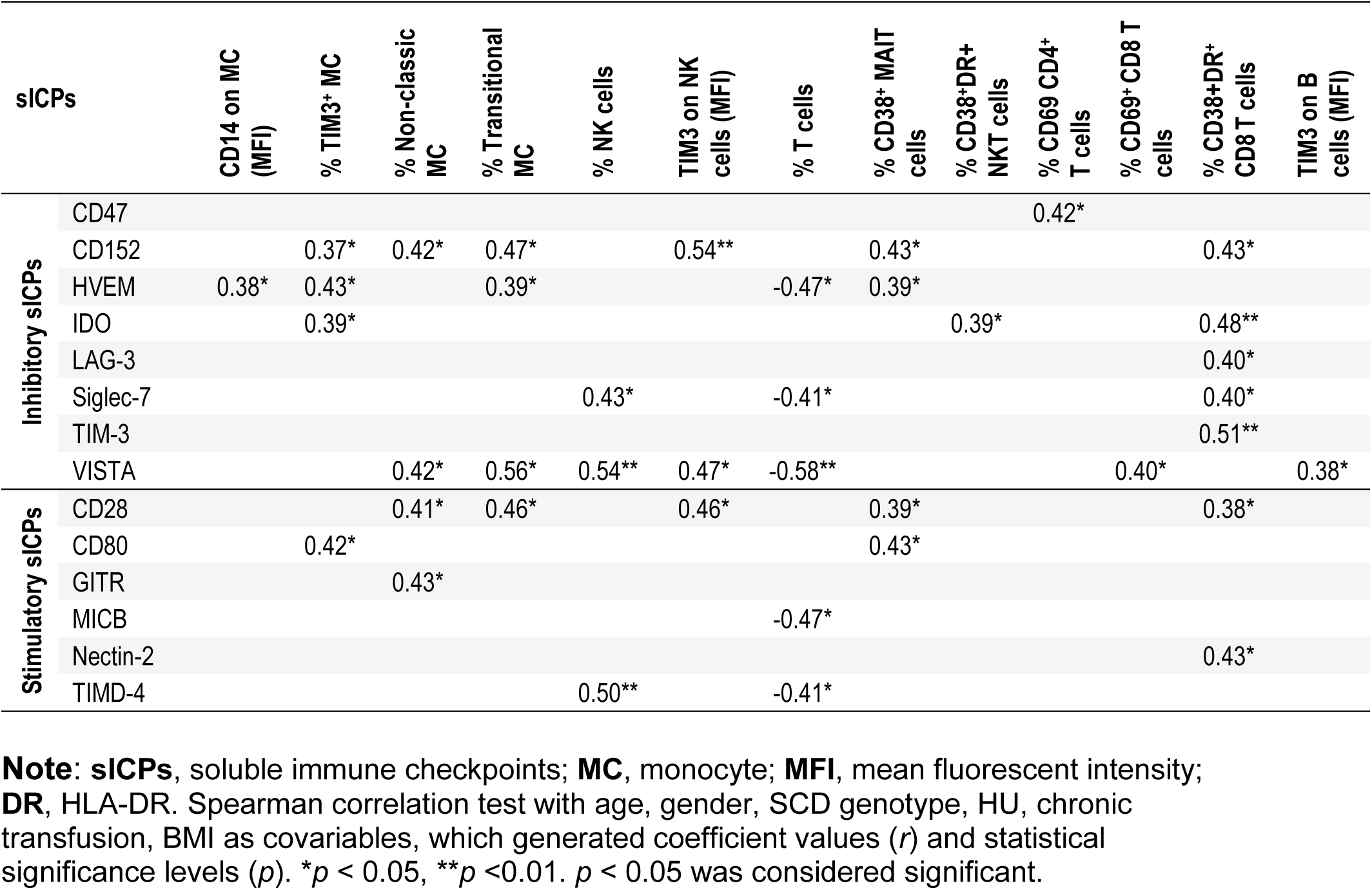
Correlations between altered plasma levels of sICPs and immune cell profiles.

### 3.5 Correlation analysis revealed associations between altered plasma levels of sICPs and PROMs

To investigate the relationship between sICPs and patient-reported outcome measures (PROMs) in SCD patients, Spearman correlation analyses were performed with and without the inclusion of covariables (Table 6 and Table S4). Covariables included age, gender, SCD genotype, HU use, BMI, and chronic transfusion status.

**Table 6.** Correlations between altered plasma levels of sICPs and PROMs.

| sICPs | PainDetect<br>Total<br>Score | FSQ<br>Widespread<br>Pain Index | PROMIS-29<br>Pain Intensity<br>Score | HADS Anxiety<br>Score | ASCQ-Me Pain<br>Episode<br>Frequency/Recency<br>Score | PedsQL<br>Total<br>Score |
| --- | --- | --- | --- | --- | --- | --- |
| **Inhibitory sICPs** |  |  |  |  |  |  |
| BTLA | -0.34<sup>&</sup> | -0.53<sup>**</sup> |  |  |  | 0.34<sup>&</sup> |
| LAG-3 |  | -0.45<sup>*</sup> | -0.47<sup>*</sup> | -0.41<sup>*</sup> |  | 0.42<sup>*</sup> |
| PD-1 | -0.39<sup>*</sup> |  |  |  | -0.36<sup>&</sup> |  |
| Siglec-7 |  |  | -0.33<sup>&</sup> |  |  | 0.35<sup>&</sup> |
| **Stimulatory sICPs** |  |  |  |  |  |  |
| CD27 |  |  |  | 0.35<sup>&</sup> |  |  |
| CD80 |  | -0.46<sup>*</sup> |  |  |  |  |

**Note:** sICPs, soluble immune checkpoints; PROMs, patient-reported outcome measures; FSQ, Fibromyalgia Survey Questionnaire; PROMIS, Patient-Reported Outcomes Measurement Information System; HADS, Hospital Anxiety and Depression Scale; ASCQ-Me, Adult Sickle Cell Quality of Life Measurement Information System; PedsQL, Pediatric Quality of Life Inventory. Spearman correlation analyses were performed including age, gender SCD genotype, HU, chronic transfusion, BMI as covariables. \* $p < 0.05$ ; \*\* $p < 0.01$ , <sup>&</sup> $p < 0.10$ , a trend toward significance.

In analyses without covariables, multiple inhibitory and stimulatory sICPs demonstrated significant correlations with multiple PROMs (Table S4). Eight (BTLA, LAG-3, PD-1, Siglec-7, CD28, CD80, CD137, and Nectin-2) had significant negative correlations with various pain measurements, while CD80 and MICA inversely correlated with the PROMIS 29 Physical Function Score. On the other hand, ten sICPs (BTLA, LAG-3, Siglec-7, TIM-3, CD80, CD134, CD137, MICA, MICB, and Nectin-2) demonstrated positive correlations with PedsQL scores.

When covariables were included, three inhibitory (BTLA, LAG-3, and PD-1) and one stimulatory (CD80) sICPs remained significantly correlated with PROMs (Table 6). Key findings include: (1) BTLA showed significant negative correlations with the PainDetect Total Score (*r* = -0.34, *p* = 0.09) and FSQ Widespread Pain Index (*r* = -0.53, *p* = 0.006), while positively correlating with the PedsQL Total Score (*r* = 0.34, *p* = 0.09), (2) LAG-3 was negatively correlated with FSQ Widespread Pain Index (*r* = -0.45, *p* = 0.02), PROMIS-29 Pain Intensity Score (*r* = -0.47, *p* = 0.01) and HADS Anxiety Score (*r* = -0.41, *p* = 0.04), but positively correlated with the PedsQL Total Score (r = 0.42, *p* = 0.03), (3) PD-1 demonstrated negative correlations with PainDetect Total Score (*r* = -0.39, *p* = 0.05)and Pain Episode (*r* = -0.36, *p* = 0.07), and (4) CD80 demonstrated negative significance in correlation with FSQ Widespread Pain Index (*r* = -0.46, *p* = 0.02).

The analyses revealed significant and intricate associations between sICPs and PROMs, which varied with the inclusion of covariables. These findings underscore the multifaceted role of sICPs in modulating pain perception and QoL in SCD patients, warranting further exploration of their immunological and clinical implications.

### 3.6 Analysis revealed associations of sICPs, inflammatory mediators, autoantibodies and immune cell phenotypes with VOCs

To identify potential underlying inflammatory traits contributing to VOCs, we performed Spearman correlation analysis of altered sICPs, inflammatory mediators, autoantibodies, and immune cell phenotypes with the time intervals from blood draw for biomarker analysis to the most recent VOC episode (Days after crisis) and to the future crisis (Days before crisis). Age, gender, SCD genotype, HU, BMI, and chronic transfusion were included as covariables. None of the sICPs and autoantibodies had significant correlations with Days after crisis and Days before crisis. As shown in Table 7, trends of negative correlations were observed between only one sICP, Nectin-2, (*r* = -0.34, *p* = 0.07) and Days post-crisis of SCD. Key inflammatory mediators, including IL-1α (*r* = -0.34, *p* = 0.07), VEGF-A (*r* = -0.34, *p* = 0.07), showed trends towards significant negative correlations with post-crisis time intervals. Notably, the frequencies of HLA-DR^+^ (*r* = -0.72, *p* = 0.0001) or CD38^+^HLA-DR^+^ (*r* = -0.48, *p* = 0.02) activated CD8 T cells were highly inversely correlated with Days after crisis. As to correlations with Days before crisis, a positive correlation for CCL23 (*r* = 0.329, *p* = 0.082) was noted, while IL-18 (*r* = -0.38, *p* = 0.04) and TIM-3 expression on B cells (*r* = -0.45, *p* = 0.03) displayed significant negative correlations.

**Table 7.** Correlations between altered plasma levels of inflammatory factors and with time interval from after and before VOC occurrence.

|  | Days after crisis |  |  |  |  | Days before crisis |  |  |
| --- | --- | --- | --- | --- | --- | --- | --- | --- |
| | Nectin-2 | IL-1α | VEGF-A | DR<sup>+</sup> CD8 T cells (%) | CD38<sup>+</sup>DR<sup>+</sup> CD8 T cells (%) | CCL23 | IL-18 | TIM3 on B cells (MFI) |
| *r* | -0.34 | -0.34 | -0.34 | -0.72 | -0.48 | 0.33 | -0.38 | -0.45 |
| *p* | 0.07 | 0.07 | 0.07 | 0.0001 | 0.02 | 0.08 | 0.04 | 0.03 |

**Note:** DR, HLA-DR; MFI, mean fluorescent intensity. The linear relationship between two variables was calculated using the Spearman correlation test with age, gender, SCD genotype, HU, chronic transfusion, BMI as covariables, which generated coefficient values and statistical significance levels ( $p$ ). $p < 0.05$ was considered significant.

Taken together, these results indicated that levels of IL-18, T cells activation, and B cell immune exhaustion were linked with VOCs and that levels of circulating IL-18 and TIM-3 expression on B cells might predict VOC occurrence.

## 4. Discussion

This study investigates the profile and role of sICPs in SCD and their associations with inflammatory mediators, autoantibodies, immune cell profiles, and clinical outcomes. Immune checkpoints (ICPs) consist of paired receptor-ligand molecules that exert inhibitory, stimulatory, or dual effects on immune regulation, surveillance, defense, and self-tolerance (23–27). ICP molecules exist in both membrane and soluble forms *in vivo* and *in vitro* (28–39). Similar to membrane-bound ICPs (mICPs), soluble ICPs (sICPs) are also present in normal physiological conditions and highly dysregulated in patients with cancer, viral infections, or alcohol-associated liver disease (ALD) (28–40). Soluble ICPs can be generated through either secretion of protein isoforms encoded by alternative mRNA splicing or protease-mediated shedding from mICPs by actions of matrix metalloproteinases (MMPs)(39, 41). Since sICPs are paired receptor-ligand molecules and circulate in the bloodstream, they likely form a circulating immune regulatory system. In addition, increasing evidence has shown that sICPs interact with their mICP compartments to positively or negatively regulate immune responses (38). Furthermore, sICPs can compete with their mICP compartments for binding to the ICP blocking antibodies, thereby interrupting the efficacy of ICP blockade therapies. Thus, there is an urgent need to study the role of sICPs in immune regulation in health and disease. Given that sICPs have not been studied in patients with SCD, we utilized clinical samples from our ongoing randomized clinical trial in SCD (ClinicalTrials.gov, NCT05045820) to study the profile and role of sICPs in SCD and their associations with inflammatory mediators, autoantibodies, immune cell profiles, and clinical outcomes. Our findings highlight the significant dysregulation of sICPs in SCD patients and their potential as biomarkers or therapeutic targets.

In comparison to HCs, SCD patients exhibited significantly elevated levels of multiple sICPs (Table 3). Among the 20 inhibitory sICPs analyzed, 7 (BTLA, CD47, CD152, HVEM, Siglec-7, TIM-3, and VISTA) were markedly elevated in SCD patients. Similarly, 8 of the 17 stimulatory sICPs (CD27, CD28, CD80, CD134, CD137, MICA, Nectin-2, and TIMD-4) showed significant increases in SCD cohort (Table 3). Of note, some ICPs exhibit dual functionality, acting as either coinhibitory or costimulatory molecules depending on the receptors or ligands they engage. For instance, HVEM (herpesvirus entry mediator, also known as TNFRSF14) is a versatile ICP that functions both as a receptor and a ligand, exerting context-dependent effects on immune regulation. As a receptor, HVEM interacts with inhibitory molecules such as BTLA (B and T lymphocyte attenuator) and CD160, promoting T-cell tolerance and maintaining immune homeostasis. These interactions inhibit T-cell receptor (TCR) signaling, recruit inhibitory signaling molecules like SHP-1 and SHP-2, and suppress cytokine production and cytotoxic activity in T cells and natural killer (NK) cells. Conversely, HVEM acts as a ligand for LIGHT (TNFSF14) and LTα (lymphotoxin-alpha), triggering immune activation. The HVEM-LIGHT axis enhances the maturation of antigen-presenting cells (APCs) and bolsters adaptive immune responses, facilitating pathogen clearance and anti-tumor immunity. Moreover, LTα-HVEM interactions contribute to lymphoid organogenesis and the formation of tertiary lymphoid structures, which play critical roles in mounting robust immune responses during infections and within tumor microenvironments. This dual functionality positions HVEM as a molecular switch, capable of toggling between immune activation and suppression based on its binding partners and the physiological context. Such properties underscore the delicate balance the immune system maintains between activation to defend against pathogens and suppression to avoid autoimmunity. Soluble HVEM has also been shown to regulate immune response, such as the production of proinflammatory cytokines IFN-γ and TNF-α by T cells and NK cells (40, 42).

Consistent with this, we found sHVEM levels in the SCD patients positively correlated with multiple inflammatory cytokines (including IFN-γ and TNF-α), chemokines, and growth factors (Fig. 1). Understanding the context-dependent roles of ICPs like HVEM in SCD is vital for addressing the chronic inflammation, immune dysregulation, and pain experienced by these patients. These insights could pave the way for targeted therapeutic strategies to modulate ICP pathways, ultimately improving clinical outcomes for individuals with SCD.

We have recently reported that inflammation and autoimmunity are interrelated in SCD patients in this study cohort (15). Following this path, we analyzed the association of sICPs with inflammatory mediators and autoantibodies. Our analysis revealed significant correlations between sICPs and various inflammatory mediators (Fig. 1). Specifically, 20 sICPs, including BTLA, CD47, CD152, HVEM, IDO, LAG-3, PD-1, Siglec-7, TIM-3, VISTA, CD27, CD28, CD80, CD134, CD137, GITR, MICA, MICB, Nectin-2, and TIMD-4, positively correlated with various proinflammatory mediators. Of note, all except MICB and Nectin-2 positively correlated with the proinflammatory cytokine IL-18, a critical proinflammatory regulator in both innate and adaptive immune responses, significantly upregulated in SCD compared to healthy controls in our recently published results (15). These findings suggest that sICPs play a crucial role in modulating inflammation in SCD. However, it is likely that inflammatory mediators reciprocally regulate sICPs and form a positive feedback loop.

Similarly, significant associations were found between sICPs and specific autoantibodies (Table 4). With covariables included, 3 sICPs (CD47, Siglec-7, and CD80), demonstrated positive correlations with autoantibodies (Table 4). Notably, the associations between sICPs and different types of autoantibodies highlight distinct pathways in autoimmunity. Siglec-7 and CD80 were significantly correlated with anti-nuclear autoantibodies (ANAs), such as anti-Sm, anti-Ribosomal P (Rib-P), and anti-Ku (Table 4), suggesting a role for these sICPs in pathways related to nuclear antigen response and immune tolerance breakdown. In contrast, CD47 showed a positive correlation with autoantibody against myeloperoxidase (MPO) (Table 4), a hallmark of small-vessel vasculitis and other systemic autoimmune diseases. This distinction between the associations of sICPs with ANAs and anti-non-nuclear autoantibodies emphasizes the diverse roles these sICPs may play in modulating specific immune responses. ANAs are primarily associated with systemic autoimmune conditions targeting nuclear components, whereas anti-non-nuclear autoantibodies are often linked to organ-specific damage and systemic inflammation. These findings provide new insights into the potential mechanistic roles of sICPs in shaping the immune landscape of autoimmune diseases, paving the way for targeted therapeutic strategies based on sICP modulation.

Both inhibitory and stimulatory sICPs demonstrated significant associations with various immune cell phenotypes, including monocytes, NK cells, T cells, MAIT cells, NKT cells, activated T cells, and B cells (Table 5). Notably, 3 sICPs (HVEM, Siglec-7, and VISTA) exhibited significant negative correlations with the percentage of T cells, but positive correlations with T cell activation (Table 5). The results suggest that sICPs are linked to the regulation of T cell dynamics and activation in the immune system of SCD patients. A significant negative correlation between sICPs and the percentage of T cells implies that higher levels of sICPs likely play a role in reducing T cell abundance, potentially by promoting T cell exhaustion, apoptosis, or suppression of T cell proliferation. These mechanisms are often observed in chronic infections (e.g., HIV) or cancer, and may also hold truth in SCD. A significant positive correlation between sICPs and T cell activation further indicates that higher levels of sICPs are associated with excessive activation and exhaustion of T cells in SCD. Alternatively, it could mean that sICPs are being released into circulation in response to T cell activation signals, serving as markers of immune system engagement.

Our analyses also demonstrated the correlations of patient-reported outcome measures (PROMs) in pain and QoL with inhibitory (BTLA, LAG-3, PD-1, and Siglec-7) and/or stimulatory (CD27 and CD80) sICPs, respectively (Table 6). Interestingly, the widespread pain index showed strong correlation with BTLA, LAG-3 and CD80, suggesting the correlation between nociplastic pain and circulating sICPs. Consistent with our previous finding (15), IL-18 is elevated prior to VOC onset (Table 7). The changes in sICPs, inflammatory mediators, autoantibodies, and immune cell phenotypes were observed in relation to SCD crisis timing (Table 7), providing insights into the immune and inflammatory dynamics surrounding VOC onset.

In conclusion, this study underscores the complex interplay between sICPs, inflammatory mediators, autoantibodies, immune cell profiles, and clinical outcomes in SCD. The significant associations between sICPs and various clinical outcome measures in pain and QoL highlight their potential as biomarkers and therapeutic targets in SCD pain management. Future research should focus on elucidating the mechanistic roles of sICPs in modulating immune activation, autoimmunity, and inflammation in SCD, which may lead to novel strategies for managing VOCs and improving the quality of life in SCD patients.

## Data Availability

All data produced in the present study are available upon reasonable request to the authors.

## 5. Author Contributions

W.L., Q.Y., and Y.W. supervised experimental performance and data collection, analysis, and interpretation; A.Q.P., L.H., C.D., and B.A.R assisted with data collection and figure preparation. W.L., Q.Y., and Y.W. drafted and edited the manuscript; Y.W. and A.R.O. directed patient recruitment; W.L. and X.H., performed statistical analyses; W.L., Q.Y., and Y.W. developed the concepts and directed experimental performance; Y.W. directed overall quality of the clinical investigation.

## 6. Abbreviations

HADS: Hospital Anxiety and Depression Scale
HC: healthy control
MFI: median fluorescent intensity
PBMC: peripheral blood mononuclear cell
PedsQL: Pediatric Quality of Life Inventory
PROMIS: Patient-Reported Outcomes Measurement Information System
ASCQ-Me: Adult Sickle Cell Quality of Life Measurement Information System
PROMs: patient-reported outcome measures
QoL: quality of life
SCD: sickle cell disease
StSt: steady-state condition
VOC: vaso-occlusive crisis.

## Acknowledgement

The authors would like to thank Tyler James Barret, Nayana Dutt, Payton Mittman, Bea Paras, Ramat Gbemisola Suleiman-Oba, Amy Gao and Yongqi Yu for assisting with experimental performance, and clinical team staff at Indiana University Clinical Research Center for patient scheduling and blood draw performance.

