## Supplementary Tables 1-4 for "Soluble immune checkpoints are dysregulated in patients with sickle cell disease and correlate with inflammatory mediators, autoantibodies, immune cell profiles, and clinical outcomes"

|  | **Table S1. Comparisons of 80 inflammatory mediators between SCD patients and healthy controls** | | | | |
| --- | --- | --- | --- | --- | --- |
|  | **Analytes (pg/ml)** | **HCs (n=40)** | **SCD (n=50)** | ***p*** | **adjusted *p*** |
| **Inflammatory cytokines** | IFN-α | 0.9 (0.9-2.1) | 1.3 (0.7-2.4) | 0.356819 | 0.998700 |
|  | IFN-γ | 1.1 (1.1-1.8) | 2.6 (1.1-4.4) | **0.000180** | **0.003736** |
|  | IL-1α | 4 (1.8-11.1) | 6 (3.8-14.7) | **0.026129** | 0.664871 |
|  | IL-1β | 0.7 (0.6-0.7) | 0.7 (0.7-1.5) | **0.044462** | 0.757545 |
|  | IL-2 | 3 (1-6.1) | 8.3 (3.5-15.8) | **0.004828** | 0.152247 |
|  | IL-3 | 5.8 (3.2-124.6) | 20.1 (3.2-107.1) | 0.289464 | 0.998700 |
|  | IL-4 | 10.1 (1.3-25.3) | 32.9 (16.6-63.5) | **0.000148** | **0.004141** |
|  | IL-5 | 1 (1-1.4) | 3 (1.2-12.1) | **0.000148** | **0.001680** |
|  | IL-6 | 1.3 (1.3-1.3) | 2.3 (1.3-7.8) | **0.001529** | **0.033864** |
|  | IL-9 | 0.7 (0.7-1) | 1.4 (0.7-3.3) | **0.016760** | 0.387632 |
|  | IL-10 | 0.4 (0.4-0.4) | 0.4 (0.4-0.4) | 0.405350 | 0.999941 |
|  | IL-12p70 | 0.7 (0.7-0.7) | 0.7 (0.7-0.7) | 0.606000 | >0.999999 |
|  | IL-13 | 0.5 (0.5-0.5) | 0.5 (0.5-0.6) | 0.189278 | 0.994058 |
|  | IL-15 | 2.8 (0.3-6.2) | 5.2 (3.6-11.3) | **0.002034** | **0.048264** |
|  | IL-16 | 129.2 (97.5-166) | 125.8 (85.2-209) | 0.606000 | >0.999999 |
|  | IL-17A | 0.4 (0.4-0.9) | 1 (0.4-3.7) | **0.045532** | 0.747497 |
|  | IL-18 | 3.6 (2.1-6) | 11.2 (5.9-41) | **0.000003** | **0.000046** |
|  | IL-21 | 17.4 (4.6-55.5) | 20.4 (13-71.7) | 0.145458 | 0.986787 |
|  | IL-22 | 1.8 (1.8-1.8) | 1.8 (1.8-2.5) | 0.606000 | >0.999999 |
|  | IL-23 | 1.5 (1.5-1.5) | 1.5 (1.5-18.7) | 0.056964 | 0.887916 |
|  | IL-27 | 1 (1-1) | 1 (1-9.5) | **0.045228** | 0.728061 |
|  | IL-31 | 1.6 (1.6-3.6) | 3.6 (1.6-22.3) | 0.056964 | 0.885507 |
|  | IL-37 | 0.3 (0.3-0.3) | 0.3 (0.3-0.3) | 0.484912 | 0.999983 |
|  | MIF | 62.3 (39.5-78.5) | 76.3 (46.7-104.4) | **0.017190** | 0.499967 |
|  | TNF-α | 2 (1.7-2.6) | 2.5 (2.1-3.2) | **0.001342** | **0.037117** |
|  | TNF-β | 0.7 (0.7-0.7) | 1 (0.7-4.5) | **0.021782** | 0.478939 |
|  | TSLP | 1 (0.6-1.9) | 2.2 (1.6-3.6) | **0.000030** | **0.000611** |
| **Chemokines** | BLC | 59.5 (37.2-109.3) | 49.5 (25.1-88.5) | 0.092214 | 0.930241 |
|  | CCL1 | 1.1 (0.7-2.7) | 1.9 (0.9-3.5) | 0.084031 | 0.887916 |
|  | CCL17 | 1.9 (1.3-2.8) | 2.3 (1.6-5.1) | 0.151877 | 0.971855 |
|  | CCL21 | 1 (1-1.5) | 21.5 (1-42.2) | **0.000824** | **0.015005** |
|  | CCL23 | 445.5 (329.5-629.2) | 892.8 (606.9-1206.7) | **<0.000001** | **0.000001** |
|  | CCL25 | 51.2 (37.1-83) | 60.5 (30.1-119.9) | 0.588324 | >0.999999 |
|  | CXCL6 | 16.2 (9.4-20.8) | 6.4 (4.9-14.8) | **0.001254** | 0.067951 |
|  | ENA-78 | 52.5 (15.1-101.2) | 75.3 (23.5-120.8) | 0.099330 | 0.927063 |
|  | Eotaxin | 7.9 (6-12.7) | 8.5 (5.6-13.8) | 0.548635 | >0.999999 |
|  | Eotaxin-2 | 0.5 (0.5-3) | 8.3 (0.5-81.6) | **0.006732** | 0.231937 |
|  | Eotaxin-3 | 0.6 (0.2-1.3) | 1.1 (0.7-1.8) | **0.010546** | 0.388438 |
|  | Fractalkine | 5.1 (1-10.1) | 4.5 (1.6-10.1) | 0.548635 | >0.999999 |
|  | GRO-α | 0.4 (0.4-1.9) | 1.9 (0.4-3) | **0.006710** | 0.208870 |
|  | IL-8 | 0.3 (0.3-4.8) | 5.5 (1.2-10.2) | **0.000251** | **0.004812** |
| **Chemokines** | IP-10 | 18.2 (11.2-24.7) | 32 (20.1-59.8) | **0.000180** | **0.005959** |
|  | I-TAC | 17.3 (6.2-37.7) | 21.3 (8.5-44.4) | 0.163265 | 0.995028 |
|  | MCP-1 | 34 (21.9-48.8) | 30.8 (15.3-49.9) | 0.168645 | 0.995924 |
|  | MCP-2 | 0.1 (0.1-0.7) | 1.8 (0.6-3.8) | **<0.000001** | **0.000001** |
|  | MCP-3 | 0.4 (0.4-2.9) | 0.9 (0.4-8.8) | 0.055997 | 0.785592 |
|  | MCP-4 | 5.7 (2.6-12.7) | 10.5 (4.3-20.7) | **0.045532** | 0.885225 |
|  | MDC/CCL22 | 54 (33-66.1) | 72 (44.4-111.1) | **0.016542** | 0.420539 |
|  | MIG | 14.2 (8.6-33.7) | 20.9 (14.2-36.1) | 0.060848 | 0.887916 |
|  | MIP-1α | 0.3 (0.3-0.3) | 2.5 (0.3-6.7) | **0.000029** | **0.000932** |
|  | MIP-1β | 24.4 (15.5-33.5) | 47.6 (29-69.3) | **0.000010** | **0.000176** |
|  | MIP-2α | 1.9 (1.3-3.3) | 2.7 (1.2-6) | 0.084725 | 0.898007 |
|  | MIP-3α | 2.5 (1.1-5.1) | 5.6 (3.9-7.4) | **0.000345** | **0.007446** |
|  | MIP-3β | 149 (87.6-255) | 182 (109-321.7) | 0.084031 | 0.898007 |
| **Growth factors** | bNGF | 0.6 (0.6-1.2) | 1.4 (0.6-3.4) | **0.003448** | 0.103840 |
|  | FGF-2 | 1 (1-1) | 1.3 (1-8.5) | **0.004684** | 0.110005 |
|  | G-CSF/CSF-3 | 10.3 (1.4-45.3) | 46 (23.6-113.3) | **0.000405** | **0.005908** |
|  | GM-CSF | 1.4 (1.4-1.4) | 1.4 (1.4-19) | **0.002381** | 0.055430 |
|  | HGF | 6.8 (3.6-14.9) | 37.6 (13.7-69.3) | **0.000001** | **0.000012** |
|  | IL-7 | 0.1 (0.1-0.1) | 0.1 (0.1-0.7) | **0.045532** | 0.747497 |
|  | IL-20 | 4.2 (2.1-7.2) | 9.4 (5.8-22.6) | **0.000189** | **0.005840** |
|  | IL-34 | 12.7 (8.5-18.5) | 23.9 (16.6-38.5) | **0.000030** | **0.000464** |
|  | LIF | 1.1 (0.5-2.6) | 2.8 (1.3-6.2) | **0.002098** | **0.040726** |
|  | M-CSF | 1.6 (1.6-1.6) | 1.6 (1.6-6.9) | 0.193121 | 0.995924 |
|  | SCF | 4.3 (1.7-8.8) | 4.3 (1.9-6.9) | 0.603852 | >0.999999 |
|  | VEGF-A | 37.3 (23.9-55.6) | 61.1 (36.6-125.1) | **0.001342** | **0.045102** |
| **Soluble receptors/proteins** | APRIL | 455.1 (212.1-1502.2) | 487.5 (173.8-1132.3) | 0.588852 | >0.999999 |
|  | BAFF | 2.9 (0.5-4.9) | 4.7 (3.5-7.4) | **0.001342** | **0.022059** |
|  | CD30 | 53 (27.6-85.4) | 67.3 (46.2-156.5) | **0.017190** | 0.582929 |
|  | CD40-Ligand | 1.2 (1.2-21.9) | 13.4 (1.2-69.7) | **0.067327** | 0.914817 |
|  | Gal-3 | 7051.8 (4288.3-10407) | 19658.1 (8552.8-39418.3) | **0.000004** | **0.000024** |
|  | IL-2R | 1239.5 (859.4-1733.5) | 1772 (982.1-2473.8) | 0.056964 | 0.885507 |
|  | MMP-1 | 56.4 (26.7-91.6) | 161.6 (31.4-266.2) | **0.008710** | 0.231937 |
|  | PTX3 | 625 (401.1-1416.1) | 2309.2 (1443.8-3893.3) | **<0.000001** | **<0.000001** |
|  | TNF-RII | 74.4 (52.3-95) | 82.4 (62.8-108.4) | 0.117571 | 0.984049 |
|  | TRAIL | 12.8 (3.8-45.6) | 11.5 (4.1-31.2) | 0.588324 | >0.999999 |
|  | TREM-1 | 56.4 (56.4-192.8) | 292.5 (108.1-1118.9) | **0.004068** | 0.102219 |
|  | Tweak | 935.4 (680.5-1357.7) | 966.5 (809-1692.9) | 0.189278 | 0.995924 |
| **SPs** | Granzyme A | 4.3 (0.5-9.9) | 11.1 (6.5-25.7) | **0.000405** | **0.007310** |
|  | Granzyme B | 10.3 (5.5-30.7) | 20.7 (11.7-40.9) | **0.017190** | 0.340085 |

**Note**: **HC**, healthy control; **SCD**, sickle cell disease; **SPs**, serine proteases. The Mann-Whitney test was used to compare differences between patients with SCD and healthy controls (HCs) with and without Holm-Šídák correction for multiple comparisons. *p* < 0.05 was considered significant.

| **Table S2. Comparisons of 18 autoantibodies between SCD patients and healthy controls** | | | | | |
| --- | --- | --- | --- | --- | --- |
| **Autoantibodies (pg/ml)** | | **HCs (n=40)** | **SCD (n=50)** | ***p*** | **Adjusted *p*** |
| **Anti-nuclear** | CENP-A | 180.7 (99-412.4) | 180.8 (110.4-581) | 0.567408 | 0.970698 |
|  | CENP-B | 65.9 (15.9-489.8) | 86.2 (19-647) | 0.559204 | 0.970698 |
|  | Ku | 270.1 (135.1-614.4) | 446.6 (192.1-1193.5) | **0.022719** | 0.275108 |
|  | Mi-2 | 604.3 (205-1668.8) | 613.5 (315-1137.4) | 0.664007 | 0.970698 |
|  | PCNA | 112 (52.6-261.3) | 117.7 (62.1-345.4) | 0.480528 | 0.970698 |
|  | PM/Scl-100 | 16 (10-21) | 23.3 (19.4-28.6) | **0.000049** | **0.000890** |
|  | Ribosomal P | 26.3 (16.5-35.4) | 41.3 (29.8-51.4) | **0.000012** | **0.000231** |
|  | RNP | 1637.2 (1103.7-2764.1) | 2001.9 (1057.4-5081.5) | 0.164521 | 0.834287 |
|  | RNP/Sm | 33.6 (19.2-156.6) | 46.1 (32.9-122.3) | **0.017308** | 0.230407 |
|  | Sci-70 | 362.5 (154.5-1392.9) | 477.8 (261.6-928.5) | 0.370296 | 0.960740 |
|  | Sm | 76.1 (27.5-324.6) | 125.4 (82.1-339) | 0.063618 | 0.514732 |
|  | SSB/La | 157.4 (85.7-301.9) | 267.5 (94.9-1579.8) | **0.048375** | 0.448446 |
|  | SSA/Ro52 | 310.8 (164-637.3) | 401 (202.8-887) | 0.247756 | 0.922869 |
|  | SSA/Ro60 | 32.5 (24.3-50.5) | 56.1 (42-72.1) | **0.000046** | **0.000878** |
| **Anti-non-nuclear** | C1q | 137.5 (70-450.8) | 202.5 (86.9-424.2) | 0.444756 | 0.970698 |
|  | β2-Glycoprotein | 50.2 (40.5-70.8) | 57.7 (41.2-95.7) | 0.259570 | 0.922869 |
|  | Myeloperoxidase | 10.2 (6.4-17.2) | 17 (11.4-24.1) | **0.000399** | **0.006758** |
|  | Proteinase 3 | 149.1 (87.4-207.9) | 202.8 (106.5-402.4) | **0.031417** | 0.339640 |

**Note:** These autoantibodies can be broadly categorized into two groups: anti-nuclear autoantibodies (ANAs) and anti-non-nuclear autoantibodies. ANAs include anti-Centromere Protein A (CENP-A), anti-Centromere Protein B (CENP-B), anti-Ku, anti-Mi-2, anti-proliferating cell nuclear antigen A (PCNA), anti-PM/Scl-100, anti-Ribosomal P, anti-Ribonucleoprotein (RNP), anti-RNP/Smith (RNP/Sm), anti-Scl-70, anti-Sm, anti-Sjögren's Syndrome-related antigen B/La (SSB/La), anti-Sjögren's Syndrome-related antigen A/Ro52 kDa (SSA/Ro52), and anti-Sjögren's Syndrome-related antigen A/Ro60 kDa (SSA/Ro60). Anti-non-nuclear autoantibodies include anti-C1q, anti-β2-glycoprotein, anti-myeloperoxidase (MPO), and anti-proteinase 3. The Mann-Whitney test was employed to compare differences between SCD patients and healthy controls (HCs), with and without Holm-Šídák correction for multiple comparisons. A *p* value of <0.05 was considered statistically significant.

**Table S3. Comparisons of immune cell phenotypes and markers between SCD patients and healthy controls**

| **Cell phenotype** | | | | **HCs (n=32)** | **SCD (n=34)** | ***p*** |
| --- | --- | --- | --- | --- | --- | --- |
| Monocytes | % |  | 10.7 (5.4-13.3) | | 11.2 (4.8-22.9) | 0.291929 |
|  | Classic | % | 86.9 (76.7-92.2) | | 83.3 (63.5-90.6) | 0.299321 |
|  |  | CD14 MFI | 7134.5 (4447.5-7927.3) | | 5094 (3937.5-6918.3) | 0.094212 |
|  |  | CD69 MFI | 1764.5 (1444.5-2279.3) | | 1931.5 (1533.3-2458.8) | 0.207788 |
|  |  | HLA-DR MFI | 17900.5 (14633.3-27640.3) | | 19073.5 (14774.8-24550.8) | 0.793897 |
|  |  | PD-1 MFI | 1018 (491.5-1944.5) | | 963 (465.3-1938.3) | 0.966967 |
|  |  | TIM3 MFI | 2466.5 (1953.3-2820.3) | | 1927 (1359-2463.5) | **0.010618** |
|  | Non-C | % | 2 (1.6-4.5) | | 0.8 (0.1-2.7) | **0.019292** |
|  | Trans | % | 0.8 (0.2-1.6) | | 0.1 (0-1) | **0.007961** |
| Lymphocytes | NK cells | % | 6.7 (4.3-12.8) | | 9.6 (6-18.8) | 0.058082 |
|  |  | CD69 % | 2.8 (2.1-3.9) | | 5.3 (4-10) | **0.000001** |
|  |  | CD38 MFI | 6664.5 (3394.5-8982.3) | | 7411.5 (4734.3-11656.3) | 0.185463 |
|  |  | DR MFI | 994.5 (712.3-1217.5) | | 936 (701.8-1135.8) | 0.764558 |
|  |  | PD-1 MFI | 311.5 (152.3-417.8) | | 263.5 (154.3-423.5) | 0.946701 |
|  |  | TIM3 MFI | 760.5 (559-961.5) | | 580 (279.3-832.3) | **0.024677** |
|  | T cells | % | 83 (79.3-90.1) | | 75.2 (60.7-87.6) | **0.010618** |
|  | MAIT  cells | % | 0.8 (0.4-1.6) | | 0.8 (0.3-1.3) | 0.434455 |
|  |  | CD69 % | 13.2 (7.2-17.4) | | 23.3 (11.5-40) | **0.000799** |
|  |  | CD38+ % | 2.8 (1.5-7.7) | | 5.8 (2.8-13.9) | 0.06072 |
|  |  | HLA-DR+ % | 2.4 (0.3-4.1) | | 1.6 (0-4.5) | 0.324188 |
|  |  | CD38+HLA-DR+ % | 0.4 (0-1) | | 0.4 (0-1.9) | 0.997384 |
|  |  | PD-1+ % | 20.7 (9.4-33.7) | | 15.1 (5.1-25.7) | 0.143106 |
|  |  | TIM3 % | 0.4 (0-0.8) | | 0.1 (0-1.1) | 0.597619 |
|  | NKT  cells | % | 3.3 (1.8-8.4) | | 2.1 (1.5-8.2) | 0.401376 |
|  |  | CD69 % | 3.7 (1.3-7) | | 11.1 (3.9-26.3) | **0.000928** |
|  |  | CD38+ % | 7.9 (5-14.5) | | 12.3 (4.7-24.7) | 0.165885 |
|  |  | HLA-DR+ % | 5.3 (3.3-8.8) | | 9.7 (5-14.3) | **0.009001** |
|  |  | CD38+HLA-DR+ % | 1.6 (0.8-2.2) | | 2.3 (1.2-5.8) | **0.022179** |
|  |  | PD-1+ % | 9.1 (3.9-20.9) | | 10.7 (3.7-15.4) | 0.997458 |
|  |  | TIM3 % | 1.9 (0.6-2.8) | | 1.3 (0.1-2) | 0.162642 |
|  | CD4 T  cells | % | 63.6 (59.1-68.5) | | 63.7 (55.3-74.4) | 0.529896 |
|  |  | CD69 % | 0.5 (0.4-0.6) | | 0.8 (0.6-1.2) | **0.000039** |
|  |  | CD38+ % | 49.8 (29.4-58) | | 36.2 (27.8-53.9) | 0.228202 |
|  |  | HLA-DR+ % | 2.4 (1.9-3.2) | | 3.2 (1.6-5.6) | 0.251352 |
|  |  | CD38+HLA-DR+ % | 0.8 (0.6-1.1) | | 1.1 (0.5-1.9) | 0.211265 |
|  |  | PD-1+ % | 8.8 (6.1-19.1) | | 9.5 (6.1-21) | 0.742761 |
|  |  | TIM3 % | 0.3 (0.3-0.5) | | 0.4 (0.2-0.5) | 0.776662 |
|  | CD8 T  cells | % | 36 (31-40.1) | | 35.2 (24.4-43.3) | 0.445813 |
|  |  | CD69 % | 1.7 (1.4-2.4) | | 4 (2.9-5.5) | **<0.000001** |
|  |  | CD38+ % | 32.3 (22.7-41.2) | | 41 (19.4-49.3) | 0.22083 |
|  |  | HLA-DR+ % | 10.3 (7.2-15.6) | | 14.6 (10.2-23.4) | **0.010117** |
|  |  | CD38+HLA-DR+ % | 2.7 (1.8-5.3) | | 6.7 (2.8-10.7) | **0.001305** |
|  |  | PD-1+ % | 9.1 (6.9-15.4) | | 11.2 (8.5-18.2) | 0.059385 |
|  |  | TIM3 % | 1 (0.6-1.8) | | 0.9 (0.5-2) | 0.880969 |
|  | B cells | % | 7.1 (5.8-9.1) | | 7 (3.3-12.6) | 0.706907 |
|  |  | CD69 % | 1.4 (1-2.6) | | 1.7 (0.9-3.6) | 0.607809 |
|  |  | CD38 MFI | 5102 (3701.8-6938.3) | | 6843 (5217.3-9117.5) | **0.010217** |
|  |  | HLA-DR MFI | 20137.5 (17720.8-23949.5) | | 21125 (16239.5-23530.8) | 0.735549 |
|  |  | PD-1 MFI | 132.5 (37.2-323.8) | | 98.4 (5.4-308.3) | 0.625838 |
|  |  | TIM3 MFI | 187.5 (95.5-422.8) | | 105.4 (-1.8-354.8) | 0.065794 |

**Note**: **Non-C**, non-classic monocytes; **Trans**, transitional monocytes; **MFI**, mean fluorescent intensity. The Mann-Whitney test was used to compare differences between SCD patients and healthy controls (HCs), with and without Holm-Šídák correction for multiple comparisons. A p value of <0.05 was considered statistically significant.

**Table S4. Correlations between altered plasma levels of sICPs and PROMs (without covariables)**

| **sICPs** | | **PainDetect Total score** | **FSQ Widespread Pain Index** | **PROMIS-29 Physical Function Score** | **PROMIS-29 Pain Intensity Score** | **ASCQ-Me Pain Episode Frequency/Recency Score** | **PedsQL Total Score** |
| --- | --- | --- | --- | --- | --- | --- | --- |
| **Inhibitory sICPs** | BTLA | -0.334* | -0.372** |  | -0.287* |  | 0.313* |
|  | LAG-3 |  | -0.421** |  | -0.547*** |  | 0.423** |
|  | PD-1 | -0.318* |  |  |  | -0.410* |  |
|  | Siglec-7 |  |  |  | -0.434** |  | 0.426** |
|  | TIM-3 |  |  |  |  |  | 0.318* |
| **Stimulatory sICPs** | CD28 |  |  |  | -0.309* |  |  |
|  | CD80 |  | -0.345* | -0.300* | -0.348** |  | 0.333* |
|  | CD134 |  |  |  |  |  | 0.450** |
|  | CD137 |  | -0.313* |  | -0.410** |  | 0.431** |
|  | GITR |  |  |  | -0.304* |  |  |
|  | MICA |  |  | -0.334* |  |  | 0.326* |
|  | MICB |  |  |  |  |  | 0.355* |
|  | Nectin-2 |  | -0.303* |  |  |  | 0.321* |

**Note: PROMs**, patient-reported outcome measures; **FSQ**, Fibromyalgia Survey Questionnaire; **PROMIS**, Patient-Reported Outcomes Measurement Information System; **ASCQ-Me**, Adult Sickle Cell Quality of Life Measurement Information System; **PedsQL**, Pediatric Quality of Life Inventory. Spearman correlation analyses were performed. ^&^*p* < 0.1, **p* < 0.05, ** *p* <0.01, *** *p* < 0.001.
