## Supplementary figures and images for "Soluble immune checkpoints are dysregulated in patients with sickle cell disease and correlate with inflammatory mediators, autoantibodies, immune cell profiles, and clinical outcomes"

### Supplementary Figure 1

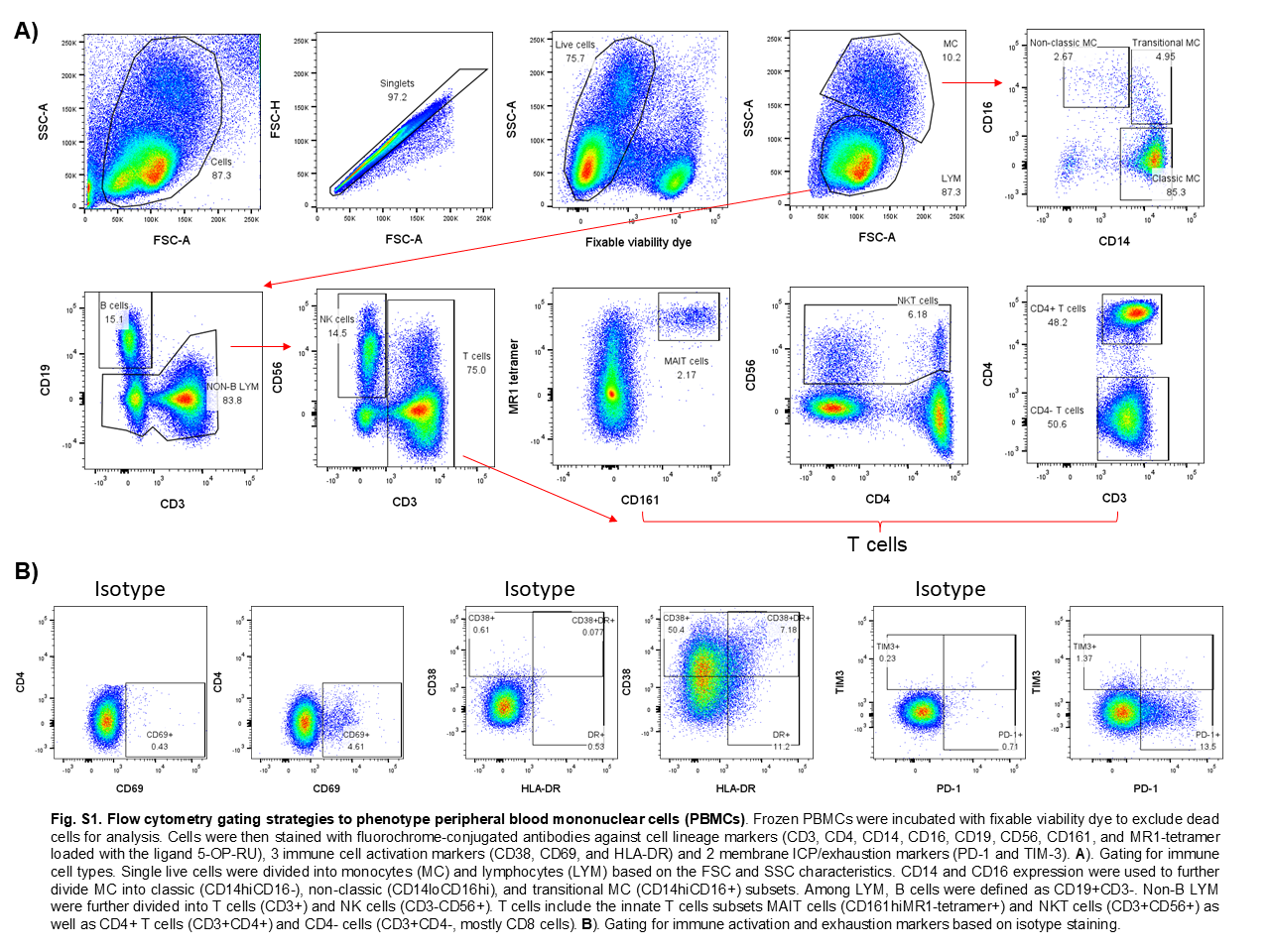
